# The Canadian platform for research online to investigate health, quality of life, cognition, behaviour, function, and caregiving in aging (CAN-PROTECT): study protocol, platform description, and preliminary analyses

**DOI:** 10.1101/2023.12.16.23300094

**Authors:** Zahinoor Ismail, Dylan Guan, Daniella Vellone, Clive Ballard, Byron Creese, Anne Corbett, Ellie Pickering, Adam Bloomfield, Adam Hampshire, Ramnik Sekhon, Pamela Roach, Eric E. Smith

**Author notes:** Corresponding Author: Zahinoor Ismail, 3280 Hospital Dr. NW Calgary AB Canada T2N 4Z6.

## Abstract

**Background:** Preventing or reducing the risk of cognitive decline and dementia is of great public health interest. Longitudinal data from diverse samples are needed to properly inform clinicians, researchers, and policy makers. CAN-PROTECT is a recently launched online observational cohort study that assesses factors contributing to both risk for incident cognitive decline and dementia and resilience against brain aging, in participants across the lifespan.

**Methods:** Measures of cognition, behaviour, and quality of life administered to both participants and study partners were compared using partial Spearman correlations adjusted for participant and study partner age, sex, and education. In participants, relationships between cognition, behaviour, function, and quality of life were examined using adjusted multivariable linear and negative binomial regression models.

**Results:** In the first three-month window, 2150 participants spanning all Canadian provinces enrolled; 637 nominated study partners had already completed assessments. Engagement with the study was excellent, with many optional assessments completed. Initial analyses demonstrated relationships between cognition, behaviour, function, and quality of life.

**Discussion:** These preliminary results speak to the utility and feasibility of CAN-PROTECT to obtain data relevant to brain health, highlighting the public interest in participating in studies on cognition. The online portal facilitated participation of a geographically diverse sample. This group is ideal to study brain aging, dementia prevention, and early detection of neurodegenerative disease. Longitudinal data will provide additional insights. Several features of CAN-PROTECT are important to consider in terms of assessing risk and resilience in Canadians, and for further development and recruitment of a research-ready cohort.

**HIGHLIGHTS:**

- CAN-PROTECT is a longitudinal online study of risk and resilience to brain aging
- Neuropsychological testing and health– and aging-related outcomes are obtained
- Data presented are from the first 2150 participants, mean age 62.9 (77.6% female)
- Associations between cognition, behaviour, function, and quality of life were found
- CAN-PROTECT is a feasible platform to obtain participant and study partner data

## INTRODUCTION

Alzheimer disease and other related dementias are devastating conditions, affecting both individuals and carers, resulting in major public health implications, and requiring substantial health care resources.[1] The emergence of disease-modifying drugs has provided some hope, however, these are complicated and labour-intensive therapies.[2] Thus, there remains great interest in prevention[3, 4], and improving our understanding of the aging brain is critical. Identifying means of preventing or reducing the risk of cognitive decline and dementia could have great benefits at individual and public health levels.[5]

While cognitive function is known to decline naturally with age, changes in memory, reasoning, or attention can be of concern to older adults and their loved ones. Indeed, age is the primary risk factor for incident cognitive decline and dementia, but there are many other potential contributors to risk.[5] Several major environmental risk factors for cognitive decline and dementia have been defined through large epidemiological and cohort studies. These factors include cardiovascular disease, depression, diabetes, dyslipidemia, hearing impairment, hypertension, obesity, and stroke[6–9]. Lifestyle factors have also been identified as significant drivers of cognitive health including physical inactivity and smoking tobacco.[8–14] More recently, the worldwide COVID-19 pandemic resulted in a great stress on older adults at risk for dementia and may have been a contributor to risk.[15]

However, personal, environmental, and lifestyle factors can also confer resilience.[16] Cognitive training[17], key dietary components like vitamin D[18] and B, antioxidants and unsaturated fatty acids[8], stimulating leisure activities, and rich social networks[19] may be protective factors. Cognitive reserve is a construct that helps operationalize the identification of these environmental and individual factors.[20] Building cognitive reserve starts in childhood (thus, dementia prevention starts in childhood too). Events across the lifespan further contribute to reserve building such as enriching childhood experiences, level and quality of education[21], complexity of occupation[22], and social and cognitive activity across the life-course.[23] Building on cognitive reserve, the scaffolding theory of aging is a multi-faceted multi-dimensional model that includes activities that build reserve, as well as those that reduce neuroplasticity.[24] In essence, this model incorporates risk and resilience, both of which are important factors to address.

How best to combine factors for risk and resilience to predict cognitive, behavioural, and functional performance in later life is a pressing issue. There remains insufficient understanding of the specific mechanisms that distinguish healthy and pathological aging. But observation and assessment of change needs to start in advance of age related cognitive and physical decline, as early as possible in the life course. Evidence that may inform clinical services, education, and public health policy needs to incorporate country-wide and region-specific factors in an ethnoculturally diverse sample, distributed across our broad geography, and collected across the lifespan. The CAN-PROTECT study aims to provide this evidence.

## METHODS

### 2.1 Study Design

CAN-PROTECT (www.can-protect.ca) is an online longitudinal observational cohort study of brain aging in community-dwelling individuals. CAN-PROTECT was developed in conjunction with the developers of the UK PROTECT study (www.protectstudy.org.uk), which is aimed at identifying factors that predict cognitive aging and dementia.[25] While there are many common elements between the two studies (e.g., neuropsychological testing, aging– and fitness-related questionnaires, physical health, lifestyle, past mental health history), CAN-PROTECT has a Canadian focus[26] and a novel assessment battery. The study includes novel assessments developed to extend the assessment of risk and resilience (e.g., new scales for quality of life, function, cognition, cognitive reserve, occupational assessment, anxious distress, and COVID-related measures, amongst others), and a unique battery of caregiver and care partner assessments (informed by experience in care settings). Collectively, the assessments were tailored to the broad geography, ethnocultural makeup, and needs of Canadian adults.

Consistent with the need to collect longitudinal data across the lifespan, all adults age ≥18 years are potentially eligible for enrolment. Participants are excluded if they have an established diagnosis of dementia or cannot provide informed consent. Participants can register and participate in the study without immediately having a study partner, although they are encouraged to find a study partner as soon as possible. Study partners must have known participants for ≥5 years. In addition to serving as a partner for a CAN-PROTECT participant, study partners can also register as CAN-PROTECT participants and name their own study partner (who may or may not be the person who named them as a study partner). Participants must be residents of Canada, have access to a computer or tablet with internet, and have a unique email address to participate in the study.

Study consent, registration, and completion of study assessments are all completed electronically. Participants and their study partners are individually asked to provide informed consent as part of the online registration process, during which they must read through a downloadable information sheet prior to checking off on the website a form containing mandatory and optional consent boxes. Optional items of the consent form include contact for newsletters and future studies, and warnings for significant drops in objective cognitive performance. Once registered, study participants first complete the demographics assessment and a cognitive test battery. Thereafter, participants can register for sub-studies, which at present comprise a caregiver study. Participants in the caregiver study are offered additional assessments about their history and experiences as current or former caregivers. Caregiver participants can be formal caregivers and care providers (e.g., paid companion, health care aid, personal care aid, or personal support worker, home care staff, licensed practical nurse, recreational therapist, occupational therapist, registered nurse, nurse practitioner, or physician) and/or informal caregivers (e.g., friend or family care partner). In total, the online registration and consent process takes approximately 20 minutes. The Conjoint Health Research Ethics Board at the University of Calgary Ethics provided approval for CAN-PROTECT (REB21-1065).

Participants are recruited from a variety of sources including local communication channels (media publicity, university press partners, online content); existing study cohorts and trials hosted by the University of Calgary; posters in primary care, geriatric, and memory clinics; strategic targeting of nation-wide seniors’ and Alzheimer disease resource centres; and social media platforms. The target sample size is >5000 participant-study partner dyads.

Participants receive ongoing communications to ensure that they complete their regular assessments and to maintain engagement in the study. These include automated assessment reminders, ad-hoc assessment notifications, regular newsletters, and access to the CAN-PROTECT YouTube channel. Participants can receive cognitive monitoring through an automated flagging system that identifies participants performing significantly lower than expected on at least two cognitive tasks on at least two separate occasions. In this scenario, the data will be examined and potentially unblinded for CAN-PROTECT study clinicians to contact flagged participants. Currently, the study is only administered in English, with plans for a French-language version in the future.

### 2.2 Study Assessments

Following registration and consent, participants receive email reminders to log into their individual study page to access the assessments. The initial assessment is for baseline demographics, after which participants gain access to the cognitive test battery, which is mandatory to continue the study. The battery consists of Trail-Making B, Switching Stroop, Self-Ordered-Search, Paired Associate Learning, Verbal Reasoning, and Digit Span tasks. With the exception of Trail-Making B and Switching Stroop, these tasks have been developed and validated in online settings.[25] Together, the battery measures cognitive domains of executive function, attention, task-switching, visual episodic memory, verbal reasoning, and working memory.[27–32] Participants must complete the cognitive test battery annually. While a complete cognitive battery comprises three test sessions in one week a minimum of 12 hours apart, only one test session is required to continue the study. There is a 50-minute time limit to complete each battery and a restriction to a one-week window to complete the cognitive battery, which are designed to optimize the quality of the cognitive test data.

After completing the cognitive test battery, participants gain access to the 16 remaining questionnaires, which they can complete in any order thy choose. Questionnaires assess subjective cognition, behaviour, function, quality of life, medical history, diet, lifestyle, physical fitness, fertility/menopause, COVID-19, family history of dementia, brain injury, mental health, and perceived health feelings and attitudes toward aging (described in detail in Table 1). Completing a cognitive battery test session also unlocks registration into sub-studies, which provides access to additional questionnaires. Formal caregivers complete a scale specific to caregiver burden in professional caregivers as well as a caregiving history and current employment status questionnaire. Informal caregivers complete a caregiver burden scale, as well as questionnaires on caregiver resources and supports and entry to role as a care provider.

**Table 1.**
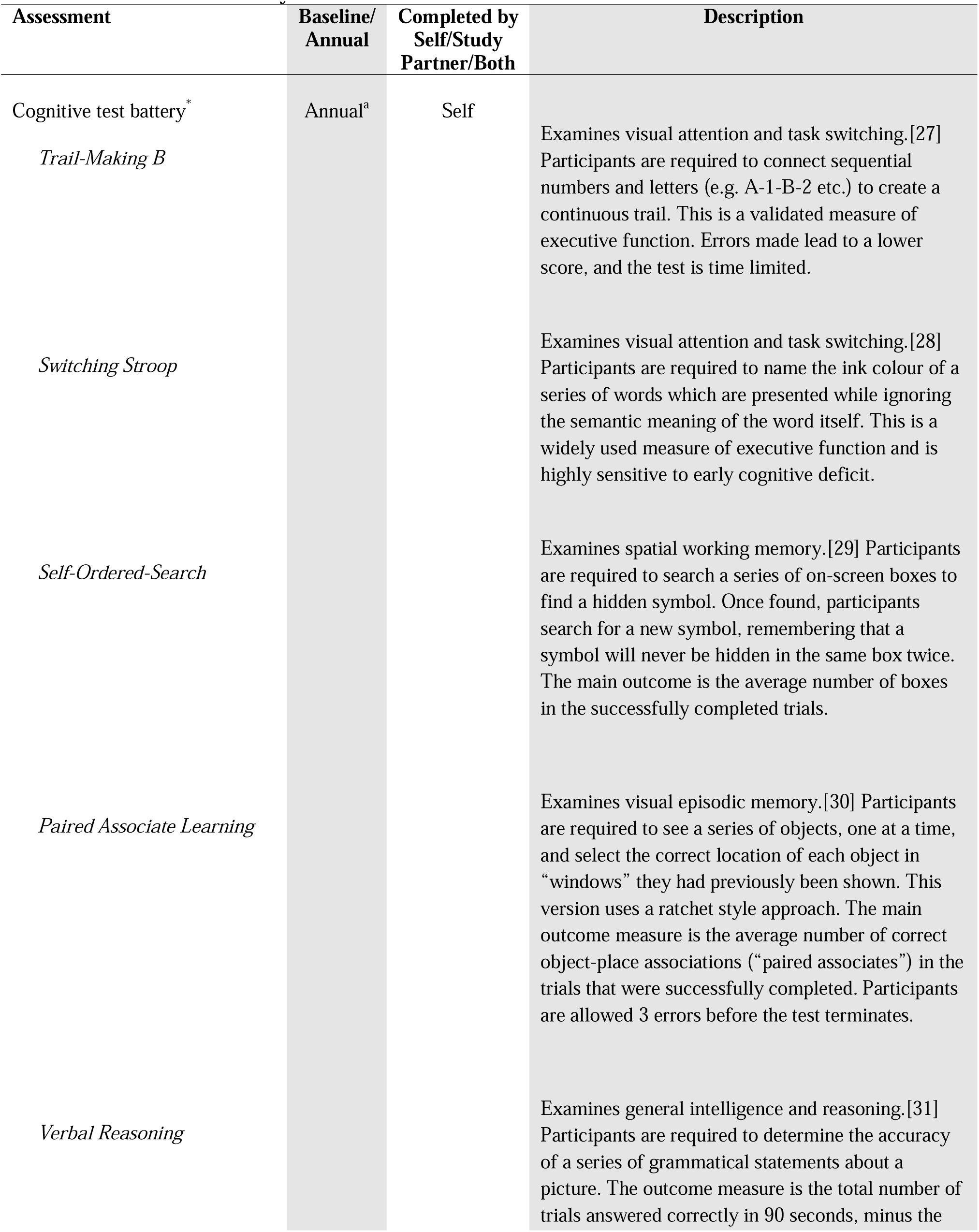
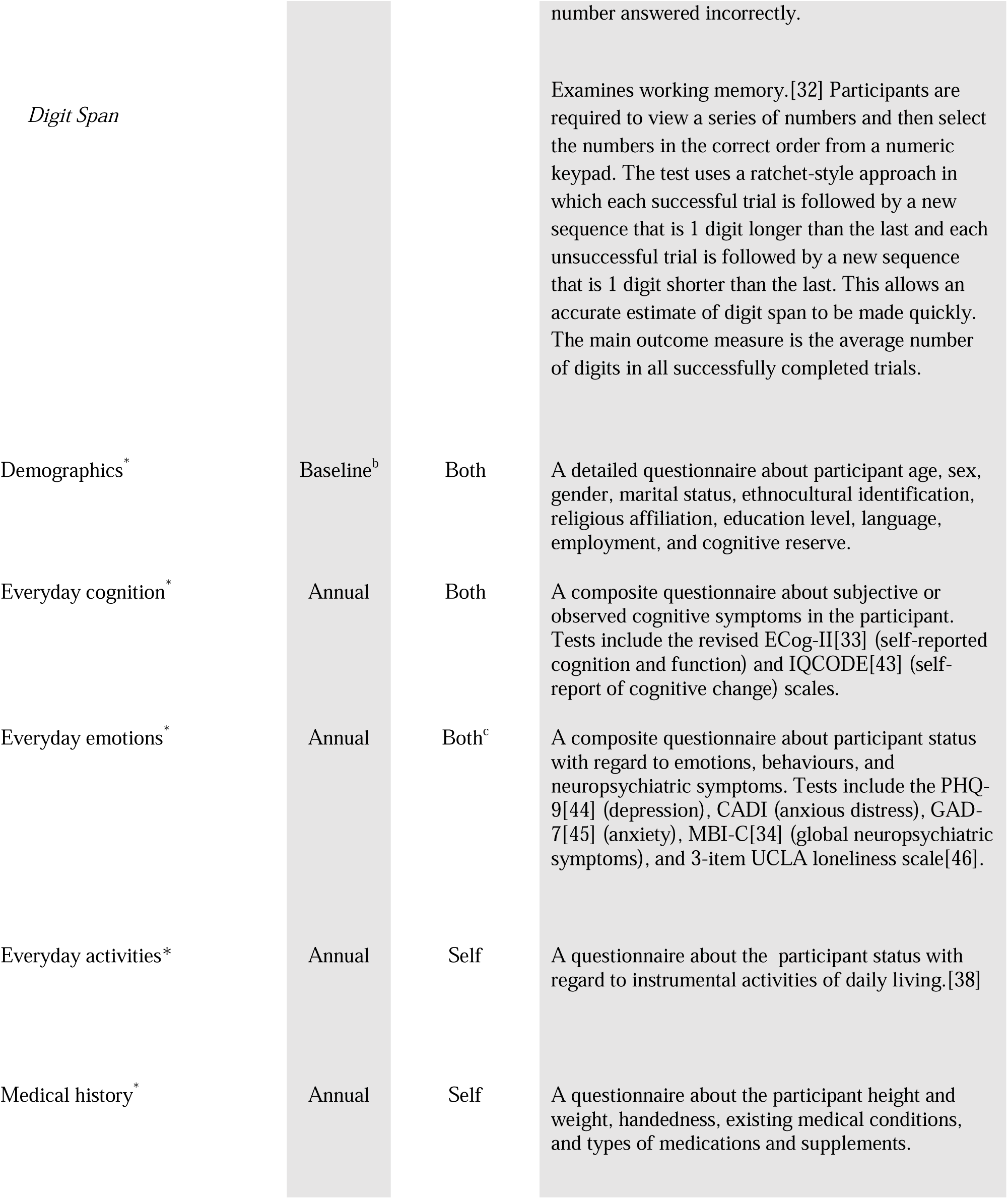
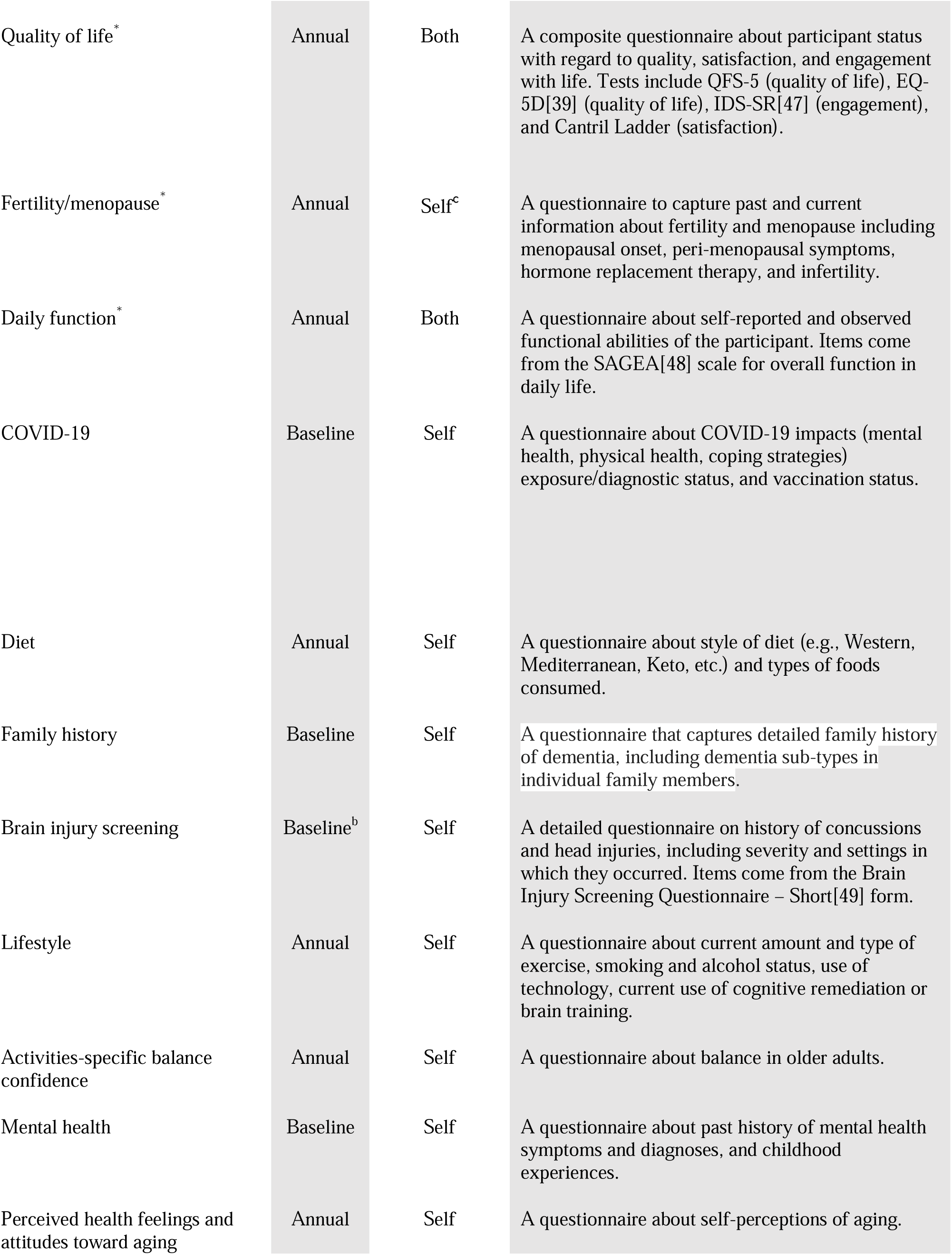
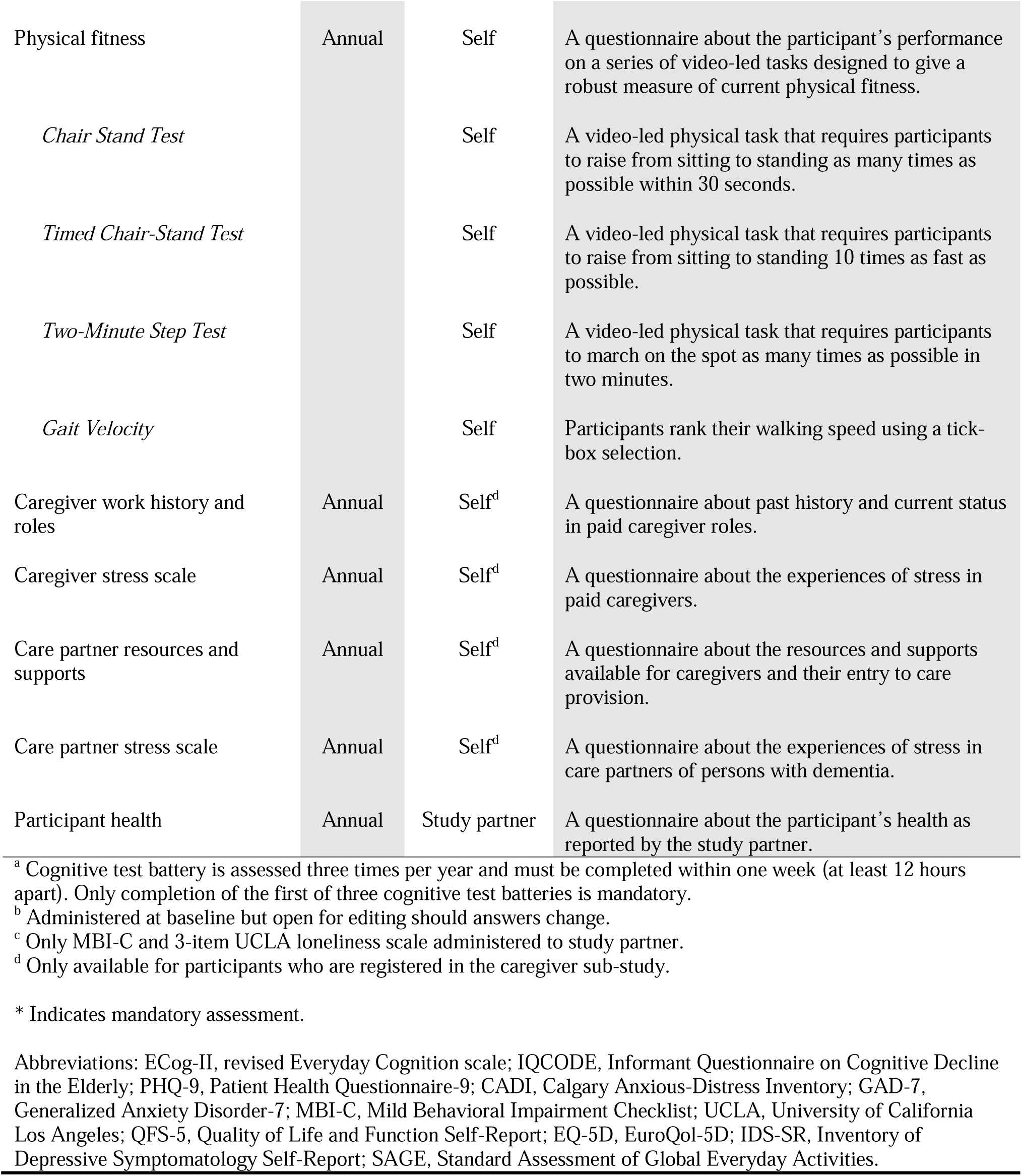
CAN-PROTECT Study Assessments.

Only 8 of the 17 participant questionnaires are mandatory, one of which is only available for female or transmen participants, allowing participants to easily tailor their involvement in the study. Furthermore, all questionnaires, can be completed over multiple sittings should the participant choose to begin but not complete them immediately. Participants are also able to freely navigate forwards and backwards across questionnaire items and revise their answers, if necessary, prior to submitting the assessment.

Annual follow-up assessments are completed through the online study platform. The annual assessment includes re-administration of the cognitive test battery, 13 of the questionnaires administered at baseline (the other four are baseline-only), and the caregiver assessments (for participants who are also caregivers). The baseline assessments take approximately 60-120 minutes, depending on how many optional assessments are completed. An additional 20 minutes is required for those completing the supplementary caregiver assessments. Annual follow-up assessments are expected to take approximately 90 minutes.

Study partners complete a questionnaire on their own demographics. Thereafter, they are offered questionnaires based on observations of participant cognition, behaviour, function, quality of life, and general health. Study partner questionnaires are similar to participant questionnaires, but are third person, often containing fewer questions than the corresponding participant questionnaires.

Participants can withdraw from the study at any time through the online study platform. In this scenario, participants may retain or destroy any personal identifiable information, but the anonymized research assessment data are retained. Participant withdrawal will also lead to automatic withdrawal of their study partner. Likewise, withdrawn study partner research assessment data are retained.

### 2.3 Data Management

All data are collected online using the custom-built CAN-PROTECT study platform. Responses to questionnaires are obtained using tick-boxes, thermometer scales, or restricted numeric data entry. Several measures have been implemented at this stage to optimize the quality of the data. These measures include numeric variables restricted to valid ranges (e.g., 18-130 years for age), skip logic for questions that are mutually exclusive (e.g., skipping questions about specific North American ethnocultural background subgroups if no North American origins are initially reported); and answer selection logic for responses that are mutually exclusive so that they cannot be simultaneously selected. All these measures were tested extensively prior to study launch across multiple individuals, devices, and browsers. Whenever a new update to the online study platform is launched, these assessments are tested extensively again to ensure high data quality and integrity.

After data collection, all personal identifiable data are stored separately from the research assessment data, with no means of linking the two datasets via the database. The research data are then anonymized prior to data extraction and analysis. Backups of the data are created daily, and the data are extracted from the database ∼monthly to ensure that users have regular access to the most up-to-date data available.

Quality control after the initial data extraction was performed manually to ensure that all variable values were coded in alignment with the detailed data dictionary and that all variable distributions were plausible. During testing, it was confirmed that the selection of specific responses to questionnaires corresponded exactly with the variable values expected based on the data dictionary. Subsequent data extractions are quality controlled via a three-step semi-automated pipeline. The first step is to determine whether any variables have been added to or removed from the dataset before evaluating all variable values against the data dictionary.

Second, statistical comparisons between corresponding variables across data extractions are performed to determine whether the distributions of any variables have deviated significantly from a previous data extraction. Third, the pipeline generates a report of the results, which includes relevant data visualizations. This report is manually reviewed by a data team member to ensure there are no unexpected deviations from previous data extractions, after which the data are released for analysis.

### 2.4 Statistical Analysis

Statistical methods for CAN-PROTECT data are tailored for each individual analysis or study question. In this initial report, we analyzed data from the initial wave of participant aged ≥40 years. We provide a descriptive statistical analysis of the data pertaining to demographics, geographic distribution, optional consents, assessment completion, as well as cognition, behaviour, function, and quality of life in study participants, caregivers, and study partners. Cognition was measured in participants according to their performance on the six objective cognitive of the battery as well as the revised Everyday Cognition (ECog-II) scale.[33] Behaviour was measured using the Mild Behavioral Impairment Checklist (MBI-C).[34, 35] Individuals were defined as having mild behavioral impairment (MBI) based on a validated MBI-C cut-off score ≥8 for dementia-free individuals.[36, 37] Function was measured using the Morris & Morris assessment for instrumental activities of daily living in older adults.[38] Quality of life was measured using the EuroQoL-5D-5L (EQ-5D).[39]

Descriptive statistics included numbers and percentages for categorical variables and means, standard deviations (SD), and ranges for numeric variables, stratified by study participants, caregivers, and study partners. Measures of cognition, behaviour, and quality of life that were administered to both participants and study partners were compared using partial Spearman correlations adjusting for participant and study partner age, sex, and education. Within participants only, the relationships between cognition, behaviour, and function were examined using multivariable linear or negative binomial regression models, as appropriate, adjusting for participant age, sex, and education.

## RESULTS

### 3.1 Demographics

In the first three months after study launch, 2150 participants ≥40 years of age (of whom 50 are caregivers) as well as 637 study partners registered in the study and had completed at least one assessment (Table 2). The mean±SD participant age was 62.9±9.3 years old and participants had completed 15.8±5.8 years of education, on average. A majority of participants reported female sex at birth (77.6%), and only three participants (0.1%) reported a non-binary or other gender. English was the most common first language of participants (92.0%), followed by Other (5.6%) and then French (2.4%). Participants tended to be Married (69.2%), although 30.8% reported a marital status other than Married (30.8%), which may include being widowed, separated, divorced, common-law, cohabitating, single, or never married. Right hand dominance was reported by 86.7% of participants, with left hand dominance in 11.0% and ambidexterity in 2.3%. Ethnocultural origins were not mutually exclusive; the most frequently reported were from Europe (83.8%), North America (49.2%), Asia (3.2%), Caribbean (1.1%), South America (0.9%), Africa (0.9%), and Oceania (0.6%). Within North America, 3.2% were First Nations, 0.1% were Inuit, 3.4% were Métis, 91.2% were Canadian, while 6.4% identified as being from the United States. Demographic characteristics of the caregiver subgroup were relatively consistent with those of the overall study participant cohort. In contrast, study partners were slightly younger (58.5±13.9 years) and more evenly distributed across sexes (54.5% female).

**Table 2.** Baseline CAN-PROTECT Participant, Caregiver, and Study Partner Characteristics.

| Variable | Participant | Caregiver | Study Partner<sup>a</sup> |
| --- | --- | --- | --- |
| n | 2150 | 50 | 637 |
| Age (years) | 62.9 (9.3), 40-91 | 61.4 (8.6), 47-81 | 58.5 (13.9), 19-87 |
| Sex |  |  |  |
| *Female* | 1669 (77.6) | 42 (84.0) | 347 (54.5) |
| *Male* | 481 (22.4) | 8 (16.0) | 290 (45.5) |
| Gender |  |  |  |
| *Woman* | 1667 (77.5) | 42 (84.0) | 346 (54.3) |
| *Man* | 480 (22.3) | 8 (16.0) | 285 (44.7) |
| *Non-binary* | 1 (0.0) | 0 (0.0) | 3 (0.5) |
| *Other* | 2 (0.1) | 0 (0.0) | 2 (0.3) |
| Education (years) | 15.8 (5.8), 0-85 | 15.8 (3.9), 6-24 | 15.6 (6.6); 1-78 |
| Language |  |  |  |
| *English* | 1979 (92.0) | 46 (92.0) |  |
| *French* | 51 (2.4) | 0 (0.0) |  |
| *Other* | 120 (5.6) | 4 (8.0) |  |
| Handedness |  |  |  |
| *Right* | 1864 (86.7) | 45 (90.0) |  |
| *Left* | 236 (11.0) | 4 (8.0) |  |
| *Ambidextrous* | 50 (2.3) | 1 (2.0) |  |
| Marital Status |  |  |  |
| *Married* | 1488 (69.2) | 39 (78.0) | 491 (77.1) |
| *Other* | 662 (30.8) | 11 (22.0) | 146 (22.9) |
| Ethnocultural Group<sup>b</sup> |  |  |  |
| *North America* | 1058 (49.2) | 23 (46.0) | 303 (47.6) |
| *First Nations* | 34 (3.2) | 0 (0.0) | 7 (2.3) |
| *Inuit* | 1 (0.1) | 0 (0.0) | 0 (0.0) |
| *Métis* | 36 (3.4) | 1 (4.3) | 11 (3.6) |
| *Canada* | 965 (91.2) | 22 (95.7) | 280 (92.4) |
| *United States* | 68 (6.4) | 1 (4.3) | 12 (4.0) |
| *Europe* | 1802 (83.8) | 41 (82.0) | 544 (85.4) |
| *Caribbean* | 23 (1.1) | 1 (2.0) | 4 (0.6) |
| *South America* | 20 (0.9) | 0 (0.0) | 4 (0.6) |
| *Africa* | 19 (0.9) | 1 (2.0) | 5 (0.8) |
| *Asia* | 68 (3.2) | 2 (4.0) | 23 (3.6) |
| *Oceania* | 13 (0.6) | 0 (0.0) | 4 (0.6) |
| Cognitive Tests<sup>c</sup> |  |  |  |
| *Trail-Making* | 62.3 (24.0), 22.1-333.1 | 54.6 (16.8), 28.9-95.5 |  |
| *Switching Stroop* | 38.1 (15.0), 0-86.0 | 42.9 (15.9), 18.0-74.0 |  |
| *Self-Ordered Search* | 6.5 (2.7), 0-13 | 6.8 (2.5), 0-9.7 |  |
| *Paired Associates Learning* | 3.9 (0.9), 0-7.3 | 4.1 (0.7), 2.0-5.3 |  |
| *Verbal Reasoning* | 31.4 (10.0), -1.5-70.0 | 35.8 (10.9), 9.0-67.0 |  |
| *Digit Span* | 6.9 (1.9), 0-20.0 | 7.2 (1.8), 2.0-11.3 |  |
| ECog-II |  |  |  |
| *Total* | 12.0 (11.5), 0-88 | 9.9 (9.5), 0-48 | 7.5 (11.7), 0-108 |
| *Memory* | 4.7 (3.9), 0-24 | 3.8 (3.1), 0-14 | 2.9 (4.0), 0-27 |
| *Language* | 3.2 (3.5), 0-24 | 2.5 (2.8), 0-13 | 1.5 (2.9), 0-27 |
| *Visuospatial Abilities* | 0.8 (1.4), 0-14 | 0.5 (0.8), 0-3 | 0.7 (1.8), 0-19 |
| *Executive Function* | 3.3 (4.6), 0-35 | 3.1 (5.0), 0-29 | 2.5 (4.4), 0-39 |
| MBI-C Score $\geq 8^d$ | 383 (26.4) | 18 (36.0) | 104 (16.3) |
| MBI-C Severity |  |  |  |
| *Global* | 5.8 (8.1), 0-65 | 7.9 (8.5), 0-41 | 4.0 (6.3), 0-45 |
| *Decreased Motivation* | 1.8 (2.7), 0-18 | 2.6 (3.1), 0-13 | 0.9 (2.0), 0-14 |
| *Affective Dysregulation* | 1.9 (2.8), 0-18 | 3.0 (2.9), 0-15 | 1.4 (2.2), 0-14 |
| *Impulse Dyscontrol* | 1.6 (2.6), 0-20 | 1.8 (2.7), 0-10 | 1.3 (2.4), 0-18 |
| *Social Inappropriateness* | 0.3 (0.8), 0-9 | 0.3 (0.7), 0-3 | 0.2 (0.6), 0-5 |
| *Psychosis* | 0.2 (0.7), 0-6 | 0.2 (0.5), 0-3 | 0.1 (0.4), 0-3 |
| IADL Total Score | 0.8 (1.8), 0-15 | 0.4 (1.2), 0-7 |  |
| EQ-5D Total Score | 6.9 (2.0), 5-19 | 7.5 (2.2), 5-15 | 7.0 (2.1), 5-20 |

<sup>a</sup> Only demographic variables pertain to the study partner. ECog-II, MBI-C, and EQ-5D values reported in the study partner column indicate scores for the participant as reported by the study partner. Some study partners may also be study participants.
<sup>b</sup> Participant and study partner are given the opportunity to identify as more than one ethnocultural group. Categories are not mutually exclusive.
<sup>c</sup> Because participants can complete the cognitive test battery up to three times per year, scores from all completed cognitive tests scores were averaged within participants before being averaged across participants to generate values reported here.
<sup>d</sup> An MBI-C total score $\geq 8$ is a validated cut-off to define older adults as having MBI.[36, 37] Because cut-off scores have not yet been validated for each MBI-C domain, prevalence rates of each MBI domain are not presented.
Numeric variables are reported in mean (standard deviation), range. Categorical variables are reported in n (%) for each level. Abbreviations: ECog-II, revised Everyday Cognition scale; MBI-C, Mild Behavioral Impairment Checklist; EQ-5D, EuroQol-5D; IADL, Instrumental activities of daily living.

The majority of participants reside in the Canadian provinces of Alberta (n=652; 30.3%), Ontario (n=562; 26.1%), and British Columbia (n=477; 22.2%). The remaining 459 (21.3%) participants are relatively evenly distributed across all other Canadian provinces. Most participants reported hearing about the CAN-PROTECT study through media publicity (n=1309; 60.9%) followed by Other (n=488; 22.7%), and word of mouth (n=273; 12.7%). Nearly all participants (96.0%) consented to receiving automated flagging system notifications for significant drops in cognitive test performance, 93.4% signed up for the study newsletters, and just over three-quarters (75.5%) of participants consented to being contacted for future studies.

### 3.2 Engagement with platform

Of the 2150 participants 1586 (73.8%) had completed the first cognitive test battery, 1064 (49.5%) the second, and 721 (33.5%) the third by the time of data extraction. Of the 1586 (73.8%) participants who completed the first cognitive test battery, which was required to access the rest of the assessments, the majority went on to complete nearly all 17 baseline participant assessments. The most frequently completed assessment was the Everyday Activities questionnaire (n=1539; 71.6%). The least frequently completed was the Perceived Health Feelings and Attitudes Toward Aging questionnaire (n=890; 41.4%), which was only applicable to participants older than 65, followed by the Physical Fitness test (n=1070; 49.8%). Among caregivers, 48 (96.0%) completed the Care Partner Stress Scale, 43 (86.0%) the Care Partner Resources and Supports questionnaire, 37 (72.0%) the Caregiver Work History and Roles questionnaire, and 5 (10.0%) the Paid Caregiver Stress Scale. Among study partners, the Everyday Emotions questionnaire was the most frequently completed assessment (n=637; 100%) and the Participant Health was the least frequently completed assessment (n=588; 92.3%).

### 3.2 Cognition, Behaviour, Function, and Quality of Life

Participants took a mean of 62.3±24.0 seconds to complete the Trail-Making B cognitive test. Average summary performance scores were 38.1±15.0 seconds for Switching Stroop, 6.5±2.7 seconds for Self-Ordered Search, 3.9±0.9 seconds for Paired Associates Learning, 31.4±10.0 seconds for Verbal Reasoning, and 6.9±1.9 for Digit Span. The average total ECog-II score was 12.0±11.5 (with higher scores denoting greater impairment). Across domains, average ECog-II scores were 4.7±3.9 for memory, 3.2±3.5 for language, 0.8±1.4 for visuospatial abilities, and 3.3±4.6 for executive function. Just over a quarter of all participants (26.4%) scored ≥8 on the MBI-C (with higher scores denoting greater symptom burden). The mean severity of MBI-C scores were 5.8±8.1 total, 1.8±2.7 for decreased motivation (apathy), 1.9±2.8 for affective dysregulation (mood/anxiety symptoms), 1.6±2.5 for impulse dyscontrol (agitation, impulsivity, abnormal reward salience), 0.3±0.8 for social inappropriateness (impaired social cognition), and 0.2±0.8 for psychosis (hallucinations and delusions). Finally, participants had an average IADL total score of 0.8±1.8 (with higher scores denoting greater impairment), and EQ-5D total score of 6.9±2.0 (with higher scores denoting poor QoL).

A total of 536 participant-study partner dyads completed assessments for cognition, behaviour, and quality of life. After controlling for participant and study partner age, sex, and completed years of education, participant and study partner reported ECog-II total scores were moderately correlated (Spearman’s r=0.30, p<.001), participant and study partner reported MBI-C total scores were weakly correlated (Spearman’s r=0.27, p<.001), and participant and study partner reported EQ-5D total scores were moderately-strongly correlated (Spearman’s r=0.53, p<.001). In participants, every 1-unit rise in MBI-C total score was associated with a 0.1 SD lower score on Trail-Making, Switching Stroop, Self-Ordered Search, Paired Associates Learning, and Digit Span (all p<0.02). MBI-C total score was not associated with any change in Verbal Reasoning performance (p=0.46). Negative binomial regressions revealed that every 1-unit rise in MBI-C total score was also associated with a 5.6% (95%CI: 4.9-6.3, p<.001) greater ECog-II total score, and 7.9% (95%CI: 6.0-9.9, p<.001) greater IADL total score.

## DISCUSSION

CAN-PROTECT is a Canada-wide online observational cohort study of brain aging. In the first three months after launch, 2150 participants ≥40 years of age from all Canadian provinces enrolled in the study; 637 of the study partners had completed assessments. Engagement with the study was excellent, with many optional assessments completed in addition to the mandatory ones. Preliminary analyses demonstrated relationships between cognition, behaviour, function, and quality of life. These results speak to the feasibility of a large-scale online study to obtain data relevant to brain aging, and to determine risk and resilience to cognitive decline. The successful launch speaks to the public interest in studies with a focus on the brain and cognition. This sample is ideal for studies of brain aging, dementia prevention, and early detection of neurodegenerative disease. Several features of CAN-PROTECT are important to consider in terms of assessing risk and resilience in Canadians, and for further development and recruitment of a potentially research-ready cohort.

### 4.1 Neuropsychological Test Battery

The neuropsychological test battery proved feasible and informative in obtaining objective information on cognitive performance across participants. Indeed, the assessment of multiple cognitive domains is essential for comprehensive studies of brain aging. CAN-PROTECT assessed the domains of attention, executive function, task switching, visual episodic memory, verbal reasoning, and working memory. These tests serve as a baseline score for each participant and can be used to determine norms for this Canada-wide sample and rates of change over time. Importantly, because assessments are online, many assessments can be obtained in a cost-effective manner, relative to conventional face-to-face testing. Further, flagging of participants with a substantial drop in cognition can identify persons who may benefit from prevention studies, both pharmacological and non-pharmacological. CAN-PROTECT has ethics approval to contact participants for potential participation in such trials if they provide consent to do so (∼75% of participants in this sample consented).

### 4.2 Geographic Reach and Diversity

Participants were recruited from all Canadian provinces. Given this broad reach of CAN-PROTECT, recruitment can further target sub-populations. Specifically, persons living in remote regions or those far from academic centres, as well as ethnocultural minorities and racialized Canadians are underrepresented in dementia research such that not enough is known about these segments of the population. Targeted enrolment of these groups can prove informative about brain aging in a more diverse population and inform future research and health services implementation.

Relatedly, the approach to identification of diversity amongst participants is a unique strength of CAN-PROTECT. The lack of participant diversity in dementia research results in speculative generalizability to more diverse real-world populations. Part of the problem is embedded in the arbitrary and vague descriptions or categorizations of ethnoracial groups in study samples. For example, the U.S. Census operationalizes ethnoracial groups by using the 1997 Office of Management and Budget Standards, which informs many studies.[40] This approach employs an archaic and reductionistic 5-class definition of race: White, Black or African American, American Indian or Alaska Native, Native Hawaiian or Other Pacific Islander, and Asian, with an “Other-specify” group (in which “multi-racial” can be written in). Two options for ethnicities are offered, Hispanic and non-Hispanic, but can only be applied to the White and Black race categories. This categorization, as often operationalized in dementia cohorts, is not sensitive with within-group variability (e.g., lumping together the monolithic Asian group) or to persons who identify and have been influenced by more than one group.

In CAN-PROTECT, the approach to ethnocultural and ethnoracial self-identification is based on a cultural mosaic model[41] and the Statistics Canada definition of ethnic origin, i.e., *the ethnic or cultural origins of the person’s ancestors*.[42] This approach allows a deeper understanding of backgrounds than the more superficial construct of race and the “melting pot” model seen elsewhere.[41] CAN-PROTECT categorizations are informed by the Canadian Census descriptions of the ethnocultural groups in Canada.[42] These categories include: 1) North American Aboriginal origins (First Nations, Inuit, Métis); 2) Other North American origins; 3) European origins (British Isles, French, Western European, Northern European, Eastern European, Southern European, Other); 4) Caribbean origins; 5) Latin, Central and South American origins; 6) African origins (Central and West African, North African, Southern and East African, Other); 7) Asian (West Central Asian and Middle Eastern, South Asian, East and Southeast Asian, Other); 8) Oceania; and 9) Pacific Islands origins. Participants can multi-select origins from drop down menus within each of the 9 categories.

Similarly, CAN-PROTECT takes a broad approach to religious affiliation, which may be an important consideration when understanding health-related decision making, and family and community inputs. Participants describe their affiliations with major Western religions, Eastern religions, Aboriginal spirituality, Agnostic and Atheist beliefs.

Sex at birth is determined, as is gender identification, including man, woman, non-binary, gender fluid, two-spirit, and other. Again, data are very sparse with respect to the roles of gender and brain aging, and the inclusion of diversity of gender affiliations will allow better assessment of gender over time. Nuanced assessments of gender may be important in understanding caregiving roles, for example.

### 4.3 Feasibility

We demonstrated the feasibility of this study as an online platform to obtain information on brain aging across the country. The high number of participants that have consented to newsletters, contact for flagging for cognitive decline, and for future studies speak to the great interest in brain aging amongst study participants. It also shows the ability for CAN-PROTECT to serve not only as a platform to observe brain aging, but also for recruitment for additional studies and trials.

### 4.4 Conclusions

CAN-PROTECT is the first Canadian study of its kind to capture near real-time assessments of a broad spectrum of adult participants, paid caregivers, and informal care partners of older adults with cognitive disorders. These data will inform the field and allow sophisticated analyses on risk and resilience in aging. This research is inclusive and represents a Canada-wide sample to optimize generalizability. Regarding caregiving, with increased recruitment the study can determine the perceived burden attendant with caregiving, the status of the caregiver/care partner, and factors which aid and exacerbate this burden. Further, paid caregivers, many of whom are immigrant women, will be well-represented in this sample, providing novel insights into this understudied group. Subsequent analyses will include participants younger than 40 years of age and explore the many novel assessments included in the study. With time, the longitudinal data will allow assessments of change, and contribute to model building for risk and resilience.

## Author Contributions

All authors contributed to critical revisions of the final manuscript as well as study design, data acquisition, and/or data analysis and interpretation. All authors provided approval for the final submission.

## Data Availability

All data produced in the present study are available upon reasonable request to the authors

